# A meta-analysis of clinically ascertained lipoedema cohorts from the UK and Spain identifies overlapping susceptibility loci with the UK Biobank

**DOI:** 10.64898/2026.02.11.26345915

**Authors:** Sara E. Dobbins, Isabel Forner-Cordero, Raquel Amigo, Laura Southgate, Katherine Hobbs, Ruby Moy, Marina Adjei, Gerard Muntané, Elisabet Vilella, Lourdes Martorell, Kristiana Gordon, Pia Ostergaard, Alan M. Pittman

**Affiliations:** School of Health & Medical Sciences, City St George’s University of London, UK; Physical Medicine and Rehabilitation Service, Hospital Universitari i Politecnic La Fe, Valencia University of Valencia, Spain; Departamento de Medicina. Universidad de Valencia, Spain; La Fe Biobank, Health Research Institute Hospital La Fe (IIS La Fe), Valencia, Spain; Oxford Brooks University, Oxford, UK; Hospital Universitari Institut Pere Mata (HUIPM), Institut d’Investigació Sanitària Pere Virgili (IISPV-CERCA), Reus, Catalonia, Spain; Departament de Medicina i Cirurgia, Universitat Rovira i Virgili, Catalonia, Spain; Centro de Investigación Biomédica en Red de Salud Mental (CIBERSAM), Instituto de Salud Carlos III, Madrid, Spain; Institut de Biologia Evolutiva (UPF-CSIC), Department of Medicine and Life Sciences, Universitat Pompeu Fabra, Parc de Recerca Biomèdica de Barcelona, Barcelona, Catalonia, Spain; Dermatology & Lymphovascular Medicine, St George’s Universities NHS Foundation trust, United Kingdom

**Author notes:** These authors contributed equally to this work (Sara E. Dobbins, Isabel Forner-Cordero, Alan M. Pittman). Correspondence to Alan Pittman.

## Abstract

Lipoedema is a chronic adipose tissue disorder mainly affecting women with excess subcutaneous fat deposition on the lower limbs, associated with pain and tenderness. There is often a family history of lipoedema, suggesting a genetic origin, but the contribution of genetics is not well studied. We conducted a genome-wide association study (GWAS) for this disorder in a clinically ascertained cohort from Spain and performed a meta-analysis with the UK lipoedema cohort GWAS. We then used the results of this study as a replication of the inferred UK Biobank “lipoedema phenotype” study.

Whilst our meta-analysis alone did not identify any genome-wide significant associations, our clinical cohorts provide support for three loci identified through the UKBB study: the chr2q24.3 *GRB14-COBLL1* locus (rs6753142, *P_META_*=1.64x10^-6^), chr6p21.1 *VEGFA* locus (rs4711750, *P_META_*=8.99x10^-7^) and the chr5q11.2 *ANKRD55-MAP3K1* locus (rs3936510, *P_META_*=1.67x10^-5^). We identify numerous rare SNPs with strong association signals in our meta-analysis (*P*<1x10^-6^) with support in both UK and Spanish datasets, three of which also show nominal support in the UKBB (*P*<0.05). These findings provide a starting point towards understanding the genetic basis of clinical lipoedema and demonstrate the utility of the interplay of large-scale biobanks genetic data and clinically ascertained cohorts to elucidate the genetic architecture of lipoedema.

## Introduction

Lipoedema is a chronic condition, predominantly affecting women, characterized by abnormal subcutaneous accumulation of adipose tissue in the limbs. The clinical phenotype is of a disproportionate figure with symmetrically enlarged lower body, typically affecting the hips and buttocks, extending to the legs, with sparing of the feet leading to a bracelet or cuffing effect at the ankles (1,2). It can be debilitating with respect to resulting pain and impaired mobility.

Recent research has provided strong evidence for a genetic basis of lipoedema (3), with estimates ranging between 40% and 89% (4–6). The genetic architecture of lipoedema is most likely complex, involving multiple genes, intricate interactions between genetic variants, and potential environmental influences, which collectively contribute to the development and progression of this disorder. GWAS studies have been performed with anthropometric measures of central obesity and fat distribution (7), identifying several genetic loci associated specifically with waist-hip ratio (WHR) independent of body mass index (BMI) and demonstrating sexual dimorphism.

A first GWAS of lipoedema was conducted on a modest cohort of patients with lipoedema (UK-lipoedema cohort), recruited from two specialist clinics in the UK (8); no locus reached genome wide significance. In 2023 Klimentidis *et al.* published a GWAS of an inferred lipoedema phenotype utilising body fat percentage and anthropometric measurement data in the UK Biobank (9), identifying 18 significant regions associated with this phenotype. Many of the genetic loci identified had also been found to associate with other features, including body fat and leptin levels. In the UK-lipoedema cohort, two significant associations also showing the same direction of effect as in UK Biobank lipoedema phenotype study (UKBB) were observed: near *VEGFA* and near *GRB14-COBLL1*.

In this study we have conducted a novel GWAS of lipoedema patients from Spain and performed a meta-analysis with the UK lipoedema cohort to identify novel genetic loci and evaluate replication of findings from the UKBB ‘lipoedema phenotype’ GWAS.

## Materials and Methods

### Spanish Lipoedema cohort ascertainment and genotyping

Patient blood samples and clinical summaries were collected through collaboration with Dr. Isabel Forner Cordero (Hospital Universitari i Politècnic la Fe, Valencia, Spain) from a lipoedema cohort (n=128) that has been previously characterized (10). Samples and data from patients included in this study were provided by the Biobanco La Fe (B.0000723) and were processed following standard operating procedures.

Patient phenotypes were checked to maintain consistency with the UK cohort (8), requiring age of onset <35yrs, BMI≤40 kg/m^2^, waist-hip ratio (WHR) ≤0.85, no or minimal android obesity, bilateral and symmetrical fat hypertrophy on lower limbs, spared feet, and persistent enlargement. Lipoedema samples were genotyped at the UCL Genomics facilities using the Illumina Infinium_Core-24_v1-2-2-a1 SNP chip. Spanish control data (n=1632) was ascertained though collaboration with Sara Bandres and had been genotyped on the Illumina NeuroChip v.1.1 SNP chip (11). Both chips broadly have the same 300,000 core, genome-wide tagging SNPs.

Genotyping data underwent quality control before association analysis using PLINK (v1.9& v2.0) (12). All samples were confirmed as female using the PLINK sexcheck function (F inbreeding coefficient, <0.2 for females). Relatedness between all sample pairs in the cohort was inferred by calculating identity by descent. In sample-pairs with PI_HAT>0.05, the sample with the highest BMI and WHR (for cases) and/or lower genotyping calling rate was excluded. A total of 28 cases were excluded from further analysis: four due to standard QC protocols, 16 relatives, six patients which did not meet our phenotypic inclusion requirements, and two due to ancestry. A total of 73 controls were excluded: 32 due to standard QC protocols, 41 due to relatedness. After standard SNP QC, selecting only SNPs genotyped on both cases and control chips, and excluding SNPs with A/T C/G alleles, 234,904 SNP remained in the analysis set.

### United Kingdom Lipoedema cohort ascertainment and genotyping

The UK samples were ascertained and genotyped as previously described (8). Briefly, patient blood samples from 148 self-reported white British were genotyped in two batches by Cambridge Genomic Services using Illumina Infinium_CoreExome-24_v1-2 single nucleotide polymorphism (SNP) chip and by UCL Genomics facilities using the Infinium_Core-24_v1-2-a1 SNP chip. 5,849 female samples of white British ethnicity enrolled in the Understanding Society UK study (https://www.understandingsociety.ac.uk/) and genotyped using HumanCoreExome-12_v1.0 were used as controls (European Genome-phenome Archive ID: EGAD00010000890). Genotyping data underwent thorough quality control as outlined above and as previously described (8).

### Association and Meta Analysis

To maximise our power to detect association signals the data was imputed as follows. SNPs with minor allele frequency (MAF) <0.05, missing call rate >0.05, and Hardy-Weinberg equilibrium ≤1x10^−6^ were excluded from the analysis. Data files were prepared utilising the Haplotype Reference Consortium (HRC) HRC-1000G-check-bim.pl tool to check for strand inconsistencies (https://www.chg.ox.ac.uk/~wrayner/tools/). Imputation was performed using the Michigan Imputation Server using SNPs passing QC common to both cases and controls. The HRC r1.1 2016 reference panel was selected, specifying the population as European and implementing Eagle for phasing. Imputation dosage files were processed using BCFtools to remove SNPs with rsq ≤ 0.3.

Association statistics for SNPs with MAF>0.01 were calculated using snptest (v2.5.6), implementing an additive frequentist test of association with the “score” method for dealing with genotype uncertainty from the imputation files. Both the UK and Spanish cohorts were processed using the same protocols as outline above. A meta-analysis of the UK and Spanish cohorts was undertaken using METAL (13)using effect size estimates and standard errors. The observed effect sizes were assessed for heterogeneity using the METAL heterogeneity command. Odd’s ratios were calculated from linear model effects estimate using approximation outlined in Cook *et al*. (14). Manhattan plots of association data were constructed using custom R scripts and ggplot2. Accounting for any population structure in SNPTEST by using the top five eigenvectors as continuous covariates in the sample file and conditioning upon these covariates, did not substantively alter the results.

### In Silico and Bioinformatics Tools

LDlink (https://ldlink.nih.gov/?tab=ldproxy) was utilised to calculate linkage disequilibrium statistics, identify proxy SNP variants, identify tagging SNPs, to search if SNPs (or variants in LD with those SNPs) have previously been associated with a trait or disease (R^2^>0.5, European Populations) in the GWAS catalog (15) and return GTEx eQTLs (https://www.gtexportal.org/home/index.html). Capture Hi-C data was extracted using the 3DGenome web portal ( https://3dgenome.fsm.northwestern.edu/) selecting “fat” as the tissue type (16,17). Plot Gardener was used for plotting of regional associations with Hi-C data and genes from UCSC hg19. Allele frequencies reported are from gnomAD v4.10 (18). CADD (19) and Alpha missense (20) were used to assess pathogenicity of missense variants.

Power calculations were performed using the GAS Power Calculator (https://csg.sph.umich.edu/abecasis/cats/gas_power_calculator/). Gene based analysis was performed using the MAGMA software (21)with: snpwise-mean model; 1000g reference data for LD calculations; and NCBI37.3 gene location data. Analysis was also performed with a window of 5kb up and downstream of gene coordinates. A threshold of *P*<2.5x10^-6^ was required for genome wide significance based on a Bonferroni correction, testing for 20,000 genes (22,23).

Approval from the appropriate Ethics and Scientific Committees Ethical was obtained from both UK (16/LO/0005) and Spanish (2011/0437 and 2014/0099) collection sites.

## Results

Following stringent quality control (Supp. Figure 1), analysis of 234,904 SNPs common to cases and controls was performed on 100 lipoedema cases and 1,559 controls from Spain. The Quantile-Quantile plot demonstrated minimal inflation of the test statistic (lambda=1.03, Supp. Figure 1), suggesting minimal impact of population stratification. The data was imputed using the HRC reference panel to over 9 million SNVs; 6,139,197 with MAF>0.01 (after imputation quality filtering, rsq≥0.8) and a meta-analysis was performed with the previously described UK-Lipoedema cohort (8) was performed. Whilst several interesting loci were identified (*P_META_*<1x10^-6^, Supp. Table 1), no significant associations were identified in the meta-analysis when requiring consistency across both studies (*P_META_*<5x10^-8^, *P_HET_*>0.05).

**Figure 1.**
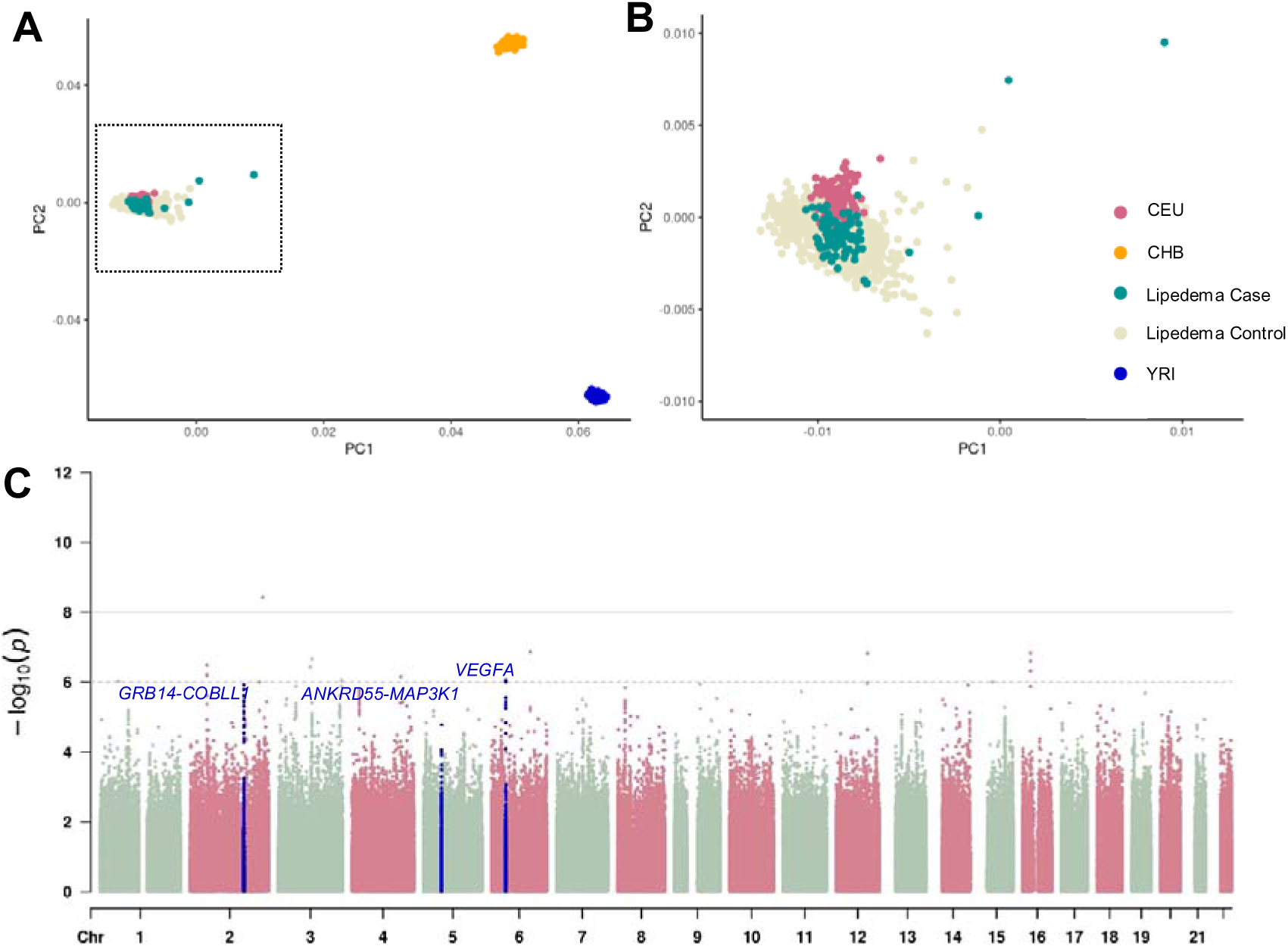
Results of the UK and Spanish genome-wide association study meta-analysis with UKBB replicated loci highlighted. (A) Plot of the first two principal components from the PCA performed on the GWAS (lipoedema cases and controls from both the UK and Spanish cohorts) and the HapMap individuals. (B) Plot of the first two principal components from the PCA zoomed in on the lipoedema cases, controls and the CEU (European ancestry) population. (C) Manhattan plot of the genome-wide *P*-values (-log_10_ scale) from the meta-analysis of the UK and Spanish cohorts. The highlighted regions demark the replicated UKBB loci (*GRB14-COBLL1, VEGFA* and *ANKRD55-MAP3K1*).

**Table 1:** Summary of significant associations from the UK Biobank GWAS with results from the UK-GWAS and Spanish-GWAS. Proxies identified: ^a^ rs7164141, ^b^rs4846567, ^c^rs28650790, **^$^**No appropriate proxy identified (R^2^>0.8). NA, no proxies identified. Odds ratios were calculated from linear model effects estimate using approximation outlined in Cook *et al*. (Cook et al., 2017) UKBB, UK biobank. Alleles: A1/A2, where A1 refers to effect allele that OR corresponds to.

|  |  |  |  | UKBB |  |  | UK |  | Spanish |  | META<br>(UK/Spain) |  |
| --- | --- | --- | --- | --- | --- | --- | --- | --- | --- | --- | --- | --- |
| UKBB top SNP | Chr:Pos | Nearest gene | Alleles | A1 Freq | OR | P value | OR | P value | OR | P value | OR | P value |
| rs72959041 | 6:127454893 | *RSPO3* | G/A | 0.95 | 1.24 | 2.70E-19 | 1.82 | 0.114 | 0.65 | 0.304 | 1.25 | 0.379 |
| rs1128249 | 2:165528624 | *GRB14-COBL1* | G/T | 0.61 | 0.92 | 3.00E-16 | 0.66 | **0.0009** | 0.68 | **0.010** | NA | **2.78E-05** |
| rs11057418 | 12:124508976 | *ZNF664-FAM101A* | G/C | 0.798 | 1.1 | 5.60E-14 | 1.19 | 0.286 | 1.19 | 0.397 | 1.18 | 0.173 |
| rs6905288 | 6:43758873 | *VEGFA* | G/A | 0.429 | 1.07 | 1.80E-13 | 1.67 | **4.45E-05** | 1.49 | **0.006** | 1.59 | **9.82E-07** |
| rs6602994<sup>a</sup> | 15:84490757 | *ADAMTSL3* | T/C | 0.276 | 0.92 | 6.90E-13 | 0.79 | 0.125 | 0.91 | 0.565 | 0.85 | 0.125 |
| rs10649697<sup>s</sup> | 5:127432908 | *SLC12A2* | T/TAGA | 0.246 | 0.92 | 1.30E-12 | NA | NA | NA | NA | NA | NA |
| rs4616635 | 3:64702275 | *ADAMTS9* | C/G | 0.723 | 0.93 | 4.90E-12 | 0.78 | 0.069 | 1.04 | 0.820 | 0.91 | 0.233 |
| rs749853052<sup>b</sup> | 1:219747226 | *LYPLAL1* | TG/T | 0.702 | 0.93 | 1.50E-11 | 0.75 | 0.028 | 0.91 | 0.545 | 0.90 | 0.040 |
| rs536569640<sup>\$</sup> | 12:123553002 | *PITPNM2* | TAATA/T | 0.726 | 1.08 | 1.90E-11 | NA | NA | NA | NA | NA | NA |
| rs28394864 | 17:47450775 | *ZNF652* | G/A | 0.538 | 1.07 | 1.90E-11 | 1.05 | 0.680 | 1.00 | 0.994 | 1.03 | 0.760 |
| rs543302184<sup>\$</sup> | 10:96009182 | *PLCE1* | T/TA | 0.58 | 1.07 | 3.00E-11 | NA | NA | NA | NA | NA | NA |
| rs11772918 | 7:46609344 | Intergenic | A/G | 0.46 | 0.93 | 3.60E-11 | 1.00 | 0.972 | 1.47 | 0.007 | 1.19 | 0.078 |
| rs62492368 | 7:150537635 | *ABP1/AOC1* | G/A | 0.694 | 1.07 | 4.00E-11 | 1.19 | 0.213 | 0.84 | 0.261 | 1.02 | 0.821 |
| rs5868014<sup>c</sup> | 5:55860907 | *ANKRD55-*<br>*MAP3K1* | GC/G | 0.814 | 1.08 | 2.70E-10 | 1.47 | **0.0326** | 1.96 | **0.002** | 1.54 | **0.0003** |
| rs71490394 | 11:62370155 | *EML3* | G/A | 0.629 | 1.07 | 1.20E-09 | 1.10 | 0.483 | 1.33 | 0.076 | 1.18 | 0.089 |
| rs28849840 | 12:50703384 | *LIMA1-FAM186A* | G/A | 0.65 | 0.94 | 1.50E-09 | 1.20 | 0.156 | 0.95 | 0.722 | 1.09 | 0.399 |
| rs565113908<sup>\$</sup> | 18:37904550 | Intergenic | T/G | 0.999 | 0.52 | 1.60E-09 | NA | NA | NA | NA | NA | NA |
| rs4680338 | 3:156794425 | *LEKR1-CCNL1* | C/G | 0.595 | 0.94 | 2.30E-09 | 0.78 | 0.046 | 0.85 | 0.249 | 0.80 | 0.023 |

Utilising biobank scale data to explore the genetic basis of lipoedema is a potentially powerful approach given the scale of samples available to analyse. However, approaches such as those by Yann *et al.* which explored a “lipoedema phenotype” in UKBB, whilst fruitful, can be difficult to interpret given the uncertain relationship between the inferred phenotype and clinically classified disease. We therefore undertook a systematic assessment of the top hits identified in the UKBB Lipoedema study (9) and identified three overlapping signals in our meta-analysis of our two clinically ascertained cohorts (Table 1, Figure 1).

The first region maps to the reported UKBB hit rs1128249 at the 2q24.3 *GRB14-COBLL1* locus (Table 1, Figure 2). Our top SNP in the region (rs6753142, chr2:165544071, *P_META_*=1.64x10^-6^) falls ∼15kb from rs1128249 (*R^2^*=0.83, *D’*=0.96) and is intronic to *COBLL1*. rs6753142 is an expression quantitative trait locus (eQTL) for *COBLL1* in thyroid and brain tissues (*P*≤0.0001), but also an eQTL for several other genes over a range of tissue types, including *SLC38A11* in adipose tissues (Supp. Table 2). Whilst our top SNP itself was not associated with any additional traits, this region harbours a wealth of GWAS signals. SNPs in LD with our top SNP are associated with a variety of traits, including anthropomorphic traits as well as metabolic biomarkers and triglyceride levels (*P*<1x10^-8^, Supp. Table 3). Missense and splice region variants in *COBLL1* have been associated with diabetic phenotypes (24). Three missense variants in weak linkage disequilibrium with our top hit (*R^2^*=0.02-0.27), all are predicted as likely benign by in-silico tools (Supp. Table 4).

**Figure 2.**
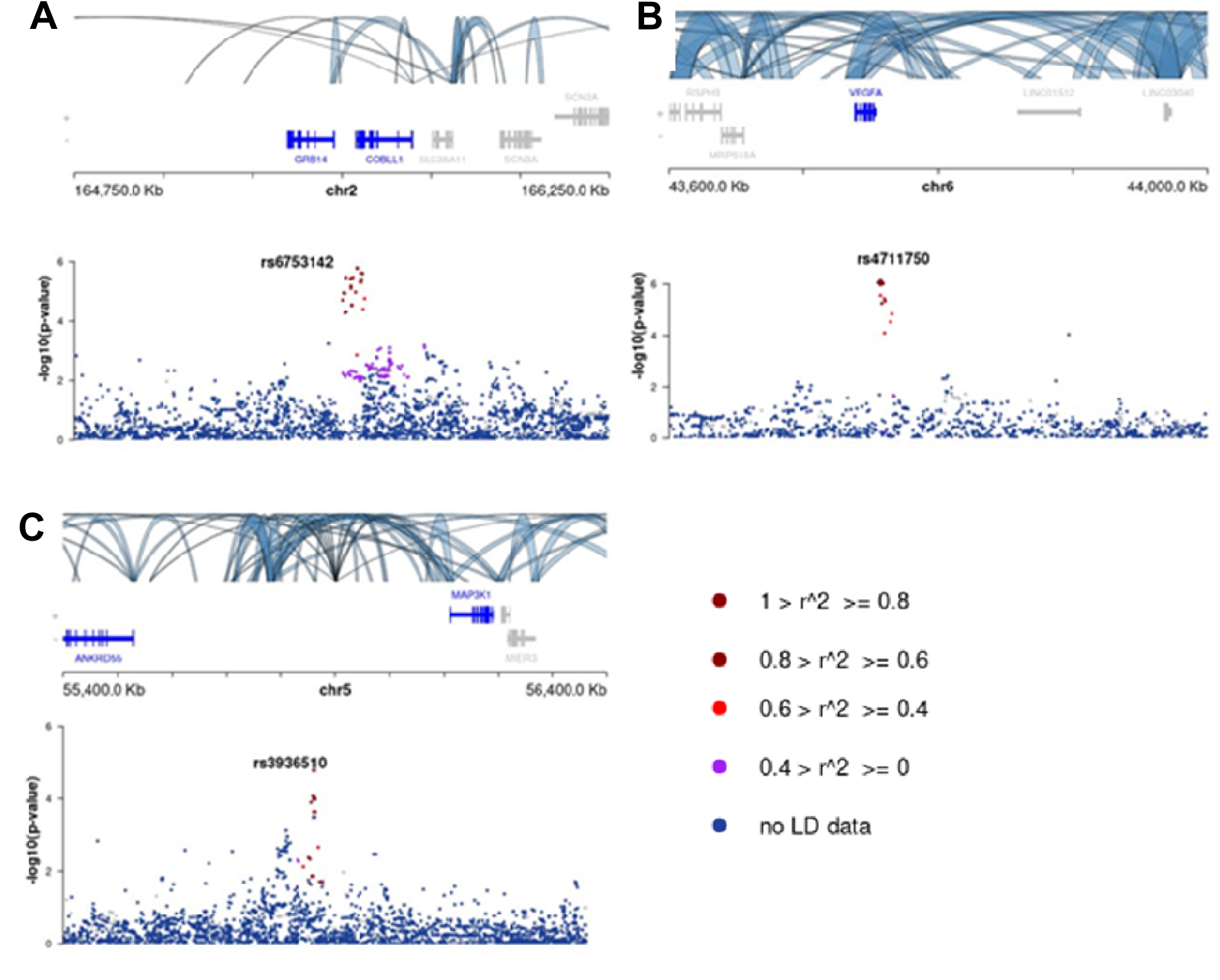
Regional association plot of UKBB regions with strong support in UK-Spanish meta-analysis. (A) Our top SNP rs6753142 (chr2:165544071) maps to the reported UKBB *GRB14*-*COBLL1* locus (rs1128249). (B) The second SNP rs4711750 (chr6:43757082) maps to the UKBB reported *VEGFA* locus. (C) The third SNP rs3936510 (chr5:55860866) maps to the *ANKRD55*-*MAP3K1* locus. The bottom panels show the P-values (in - log_10_ scale) and is colour coded by *r^2^*values with the lead SNP (labelled). The upper panels show the promoter capture Hi-C data (in fat tissue), indicating potential regulatory roles of non-coding variants with nearby genes.

The second region maps to the reported UKBB 6p21.1 *VEGFA* locus with strong support in our meta-analysis (rs4711750, chr6:43757082, *P_META_*=8.99x10^-7^, Table 1, Figure 2). This SNP, 1.8kb from the UKBB lead SNP rs6905288 (r*^2^*=0.65, *D’*=0.94) and 3kb downstream of *VEGFA*, is associated with 26 unique traits including lipid levels and metabolic markers for liver function and reticulocyte count (*P*<1x10^-8^, Supp. Table 5).

The third region with support in our meta-analysis maps to the 5q11.2 *ANKRD55-MAP3K1* locus (rs3936510, chr5:55860866, *P_META_*=1.67x10^-5^, Table 1, Figure 2), 42bp from the UKBB top hit rs5868014 (*r^2^*=0.90, *D’*=1.0). Our top SNP is associated with 35 unique traits, including anthropomorphic traits as well as type 2 diabetes and cholesterol levels (*P*<1x10^-8^, Supp. Table 6).

Despite only the 2q24.3 locus being identified as an eQTL, capture Hi-C data suggests potential regulatory roles of the nearby genes in adipose tissue for all three replicated SNPs (Figure 2).

### Loci from the UK-Spanish meta-analysis implicated in lipoedema aetiology with support from the UKBB lipoedema phenotype study

We identified an additional 16 SNPs from 12 loci with *P_META_*<1x10^-6^, with consistent direction of effect and support in both studies (Supp. Table 1). Three of these loci show support from UKBB (*P_UKBB_*<0.05).

We identify one genome-wide significant association in our meta-analysis mapping to chr2q36.1, ∼200kb distal to *KCNE4* (SNP rs79841817, chr2:224137105, *P_meta_*=3.7x10^-9^, Supp. Table 1, Supp. Figure 2). Whilst both studies support the association (*P_uk_*=0.005, *P_Spain_*=1.0x10^-9^), significant heterogeneity exists (*P_HET_*=0.001) with the effect size being considerably larger in the Spanish versus UK study. The SNP is also nominally significant in the UKBB data (*P_UKBB_*=5.2x10^-3^). However, whilst variability between studies does not preclude the possibility of a true effect, this factor, in combination with the low MAF (gnomAD total allele frequency = 0.019, v4.1.0) and lack of additional support in the region, requires caution in the interpretation of the result.

The top SNP from the association signal at 6q22.31 (rs143095903, chr6:119353044, *P_META_*=1.4x10^-7^, Supp. Table 1, Supp. Figure 2) and is intronic to *FAM184A*. Whilst no eQTLs are reported for this SNP, numerous exist for SNPS in LD including rs117839551 (*r^2^*=0.43, *D’*=1) with *RP11* in thyroid tissue (*P*=1x10^-5^) and *FAM184A* in muscle, adipose and skin tissue (*P*=1x10^-4^, *P*=2x10^-4^ and *P*=3x10^-4^ respectively, Supp. Table 7).

The association signal at chr3q13.11 (rs145150219, chr3:103862449, *P_META_*=2.2x10^-7^, Supp. Table 1, Supp. Figure 2) is located ∼1MB downstream of *ALCAM* and *CBLB*. Hi-C data demonstrates interactions in the region with promoters of *CBLB* and *ALCAM*, suggesting a potential role in the regulation of these genes, with *CBLB* interactions present only when considering fat tissue (Supp. Figure 2). *CBLB* encodes an E3 ubiquitin protein ligase with models demonstrating a potential role in obesity-induced insulin resistance in mice (25).

A gene based meta-analysis (utilising MAGMA) identified no genes as enriched after adjusting for multiple testing.

## Discussion

The meta-analysis of our UK and Spanish cohorts provides a robust replication for three of the findings from the UKBB inferred lipoedema phenotype GWAS, underscoring the relevance of these findings to clinically defined lipoedema.

VEGFA (Vascular Endothelial Growth Factor A) induces proliferation and migration of vascular endothelial cells, and whilst the relationship between lipoedema and lymphatic dysfunction is debated (26) this may suggest a link between lipoedema and vascular development. A study utilising UKBB MRI images to define “obesity axes” identified an association between the *COBLL1* locus and both peripheral and lower-body subcutaneous fat distribution (or axis) (27). Furthermore, *COBLL1* has been shown to be enriched in adipose tissue as well as demonstrating a lipodystrophy-like phenotype in *Cobll1* knock out mice (28). Understanding the biological basis of the *ANKRD55-MAP3K1* locus may be more complex, however, both *ANKRD55* and *MAP3K1* knock out cell lines show defects in preadipocyte proliferation and/or differentiation (29,30).

The meta-analysis also identifies an additional 12 loci with *P_META_*<1x10^-6^, only one of which is driven by common variation (MAF>0.05, Supp. Table 1). Given our modest cohort size, caution must be applied when interpreting these results. However, three of these loci, where the association is driven by rare variants, show some support in the UKBB and warrant further investigation. The top hit from the previous UK-lipoedema GWAS, at the *LHFPL6* locus (rs1409440, chr13:39685567 C>T; OR= 0.49, P_UK_-value 1.5x10^-5^), did not show a strong effect in the Spanish-lipoedema GWAS (OR= 0.70, P-value =0.10; *P_META_*= 1.3x10^-5^) or UKBB (P_UKBB_=0.91) although the direction of effect was consistent. Analysis of additional cohorts will be ncessary to resolve these uncertainties and provide additional power to facilitate subtype analysis.

In conclusion our study robustly supports clinically relevant associations with a lipoedema phenotype at the *GRB14-COBLL1*, *VEGFA* and *ANKRD55-MAP3K1* loci, highlighting the power of combined population based and clinically characterised cohorts for studying the genetics of lipoedema. Whilst several novel associations with rare variants show good support in all three cohorts, more data is required to validate these signals. The expansion of cohorts will be critical to further deciphering the underlying genetics of lipoedema, which should provide much needed insights into the biological basis of the condition, helping with early diagnosis and providing opportunities for therapeutic interventions.

## Data Availability

All data produced in the present study are available upon reasonable request to authors. The data will be made available via FigShare upon publication.

## Acknowledgements

The authors thank all participants for volunteering their time for this study. We would also like to thank ‘Lipoedema UK’ for facilitating recruitment through their members. Understanding Society is under the scientific leadership of the Institute for Social and Economic Research (ISER), University of Essex, and survey delivery by NatCen Social Research and Kantar Public. The research data are distributed by the UK Data Service. We also extend our thanks to members of the City St George’s, University of London (CSGUL) Lymphovascular Research Team for invaluable discussions and feedback on our work.

## Supporting Information Captions

**Supplementary Table 1**: Top hits from META analysis between UK and Spanish cohort

**Supplementary Table 2:** eQTL GRB14-COBBL locus from ldlink

**Supplementary Table 3:** GWAS catlog results for GRB14-COBBL locus top SNP

**Supplementary Table 4:** Coding SNPs in LD with rs6753142

**Supplementary Table 5:** GWAS catlog results VEGFA locus

**Supplementary Table 6:** GWAS catlog results forANKRD55-MAP3K1 locus

**Supplementary Table 7:** eQTL FAM184A locus from ldlink

**Supplementary Figure 1** : Quantile–Quantile plot of Spanish GWAS (cases and controls) showing no significant genomic inflation (lambda=1.03).

**Supplementary Figure 2: Regional association plots of top three novel regions from meta-analysis with support in UKBB.** (A) Top region maps to chr2q36.1 (lead SNP rs79841817) ∼200kb distal to KCNE4. (B) Top SNP rs143095903 from the association signal at 6q22.31 is intronic to FAM184A. (C) The third region map to chr3q13.11 (lead SNP rs145150219) downstream of ALCAM and CBLB. The bottom panels show the P-values (in - log_10_ scale) and is colour coded by *r^2^* values with the lead SNP (labelled). The upper panels show the promoter capture Hi-C data (in fat tissue), indicating potential regulatory roles of non-coding variants with nearby genes.□□

## Notes

### Competing Interest Statement

The authors have declared no competing interest.

### Funding Statement

This project was supported by Lipedema Foundation (https://www.lipedema.org/) LF36_21 (AMP, KG, SED and PO).
The study was supported by the Catalan Agency of Research and Universities (AGAUR, grant number 2021SGR01065, EV, GM, LM). https://agaur.gencat.cat/en/lagaur/qui-som/index.html

### Author Declarations

Ethics committee of London - Fulham approval gave ethical approval for this work (16/LO/0005) Research Ethics Committee of the La Fe University and Polytechnic Hospital gave ethical approval for this work (2011/0437 and 2014/0099)

