## Supplementary Figures 1-2 for "A meta-analysis of clinically ascertained lipoedema cohorts from the UK and Spain identifies overlapping susceptibility loci with the UK Biobank"

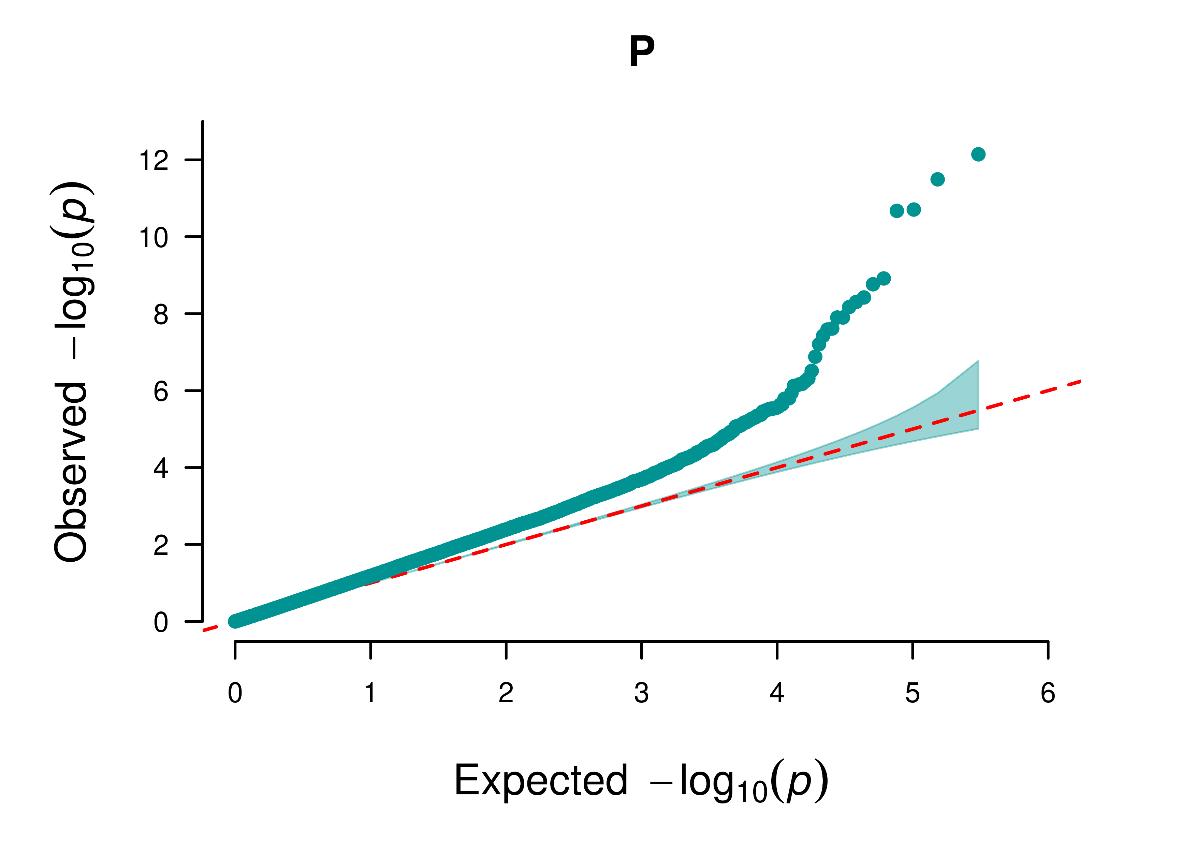

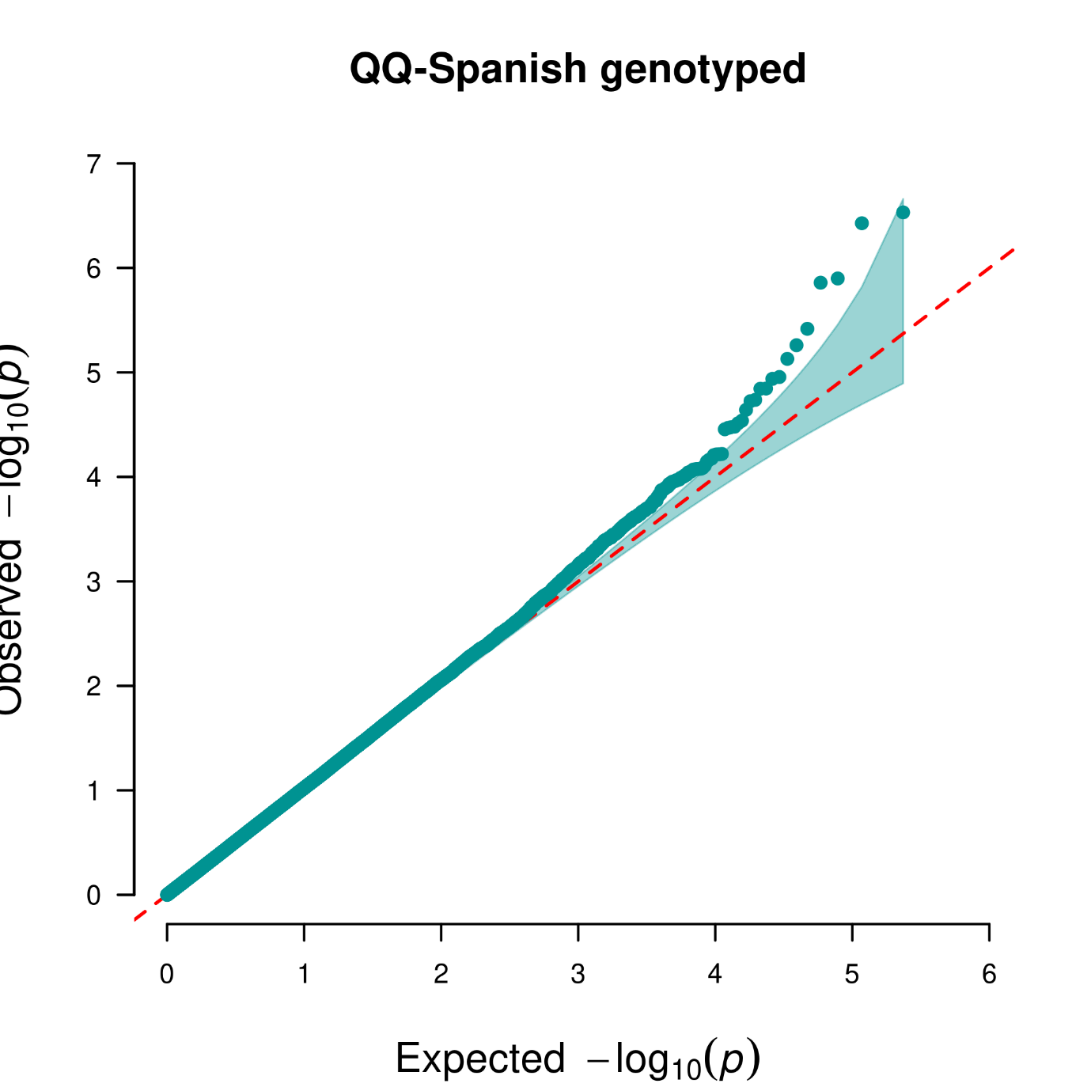

**Supplementary Figure 1**

Quantile–Quantile plot of Spanish GWAS (cases and controls) showing no significant genomic inflation (lambda=1.03).

**A**

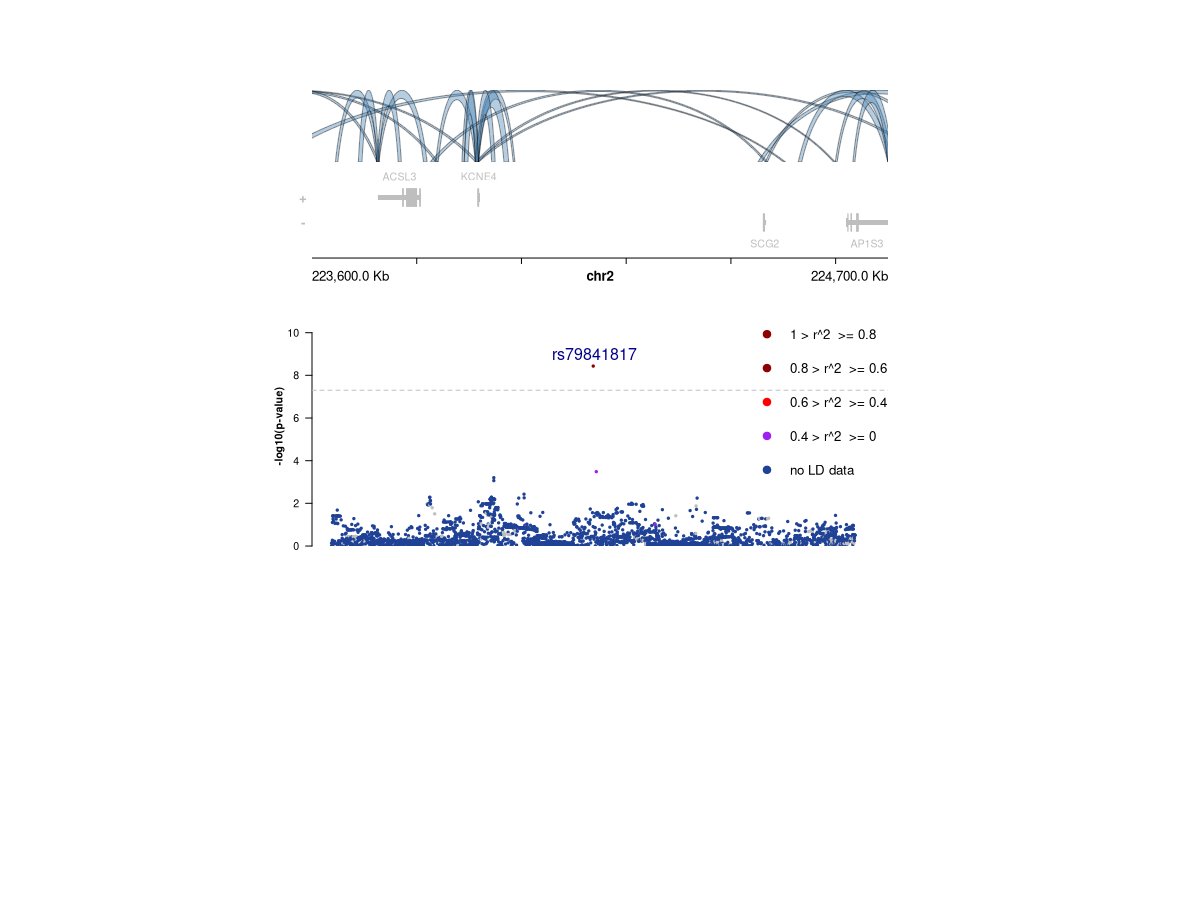

**B**

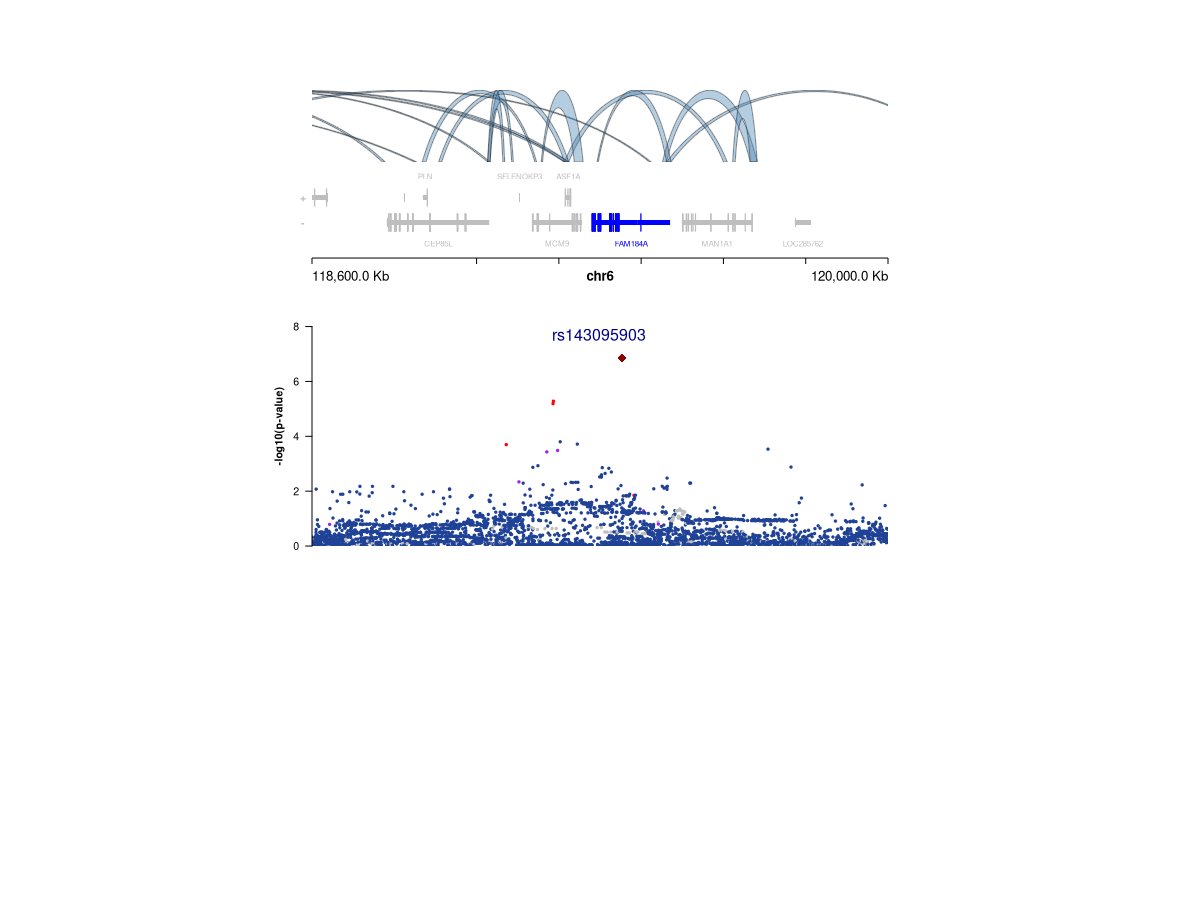

**C**

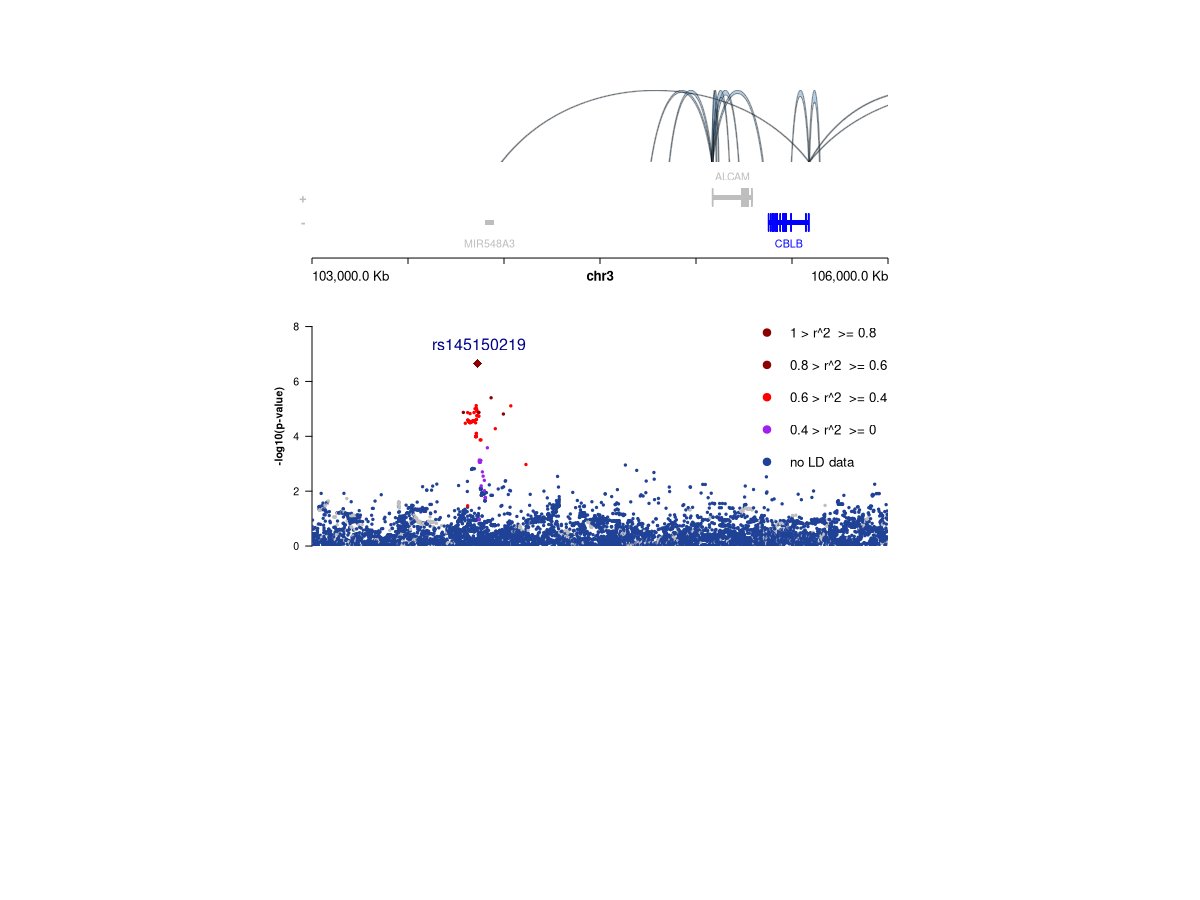

**Supplementary Figure 2: Regional association plots of top three novel regions from meta-analysis with support in UKBB.**

(A) Top region maps to chr2q36.1 (lead SNP rs79841817) ~200kb distal to KCNE4. (B) Top SNP rs143095903 from the association signal at 6q22.31 is intronic to FAM184A. (C) The third region map to chr3q13.11 (lead SNP rs145150219) downstream of ALCAM and CBLB. The bottom panels show the P-values (in -log_10_ scale) and is colour coded by *r^2^* values with the lead SNP (labelled). The upper panels show the promoter capture Hi-C data (in fat tissue), indicating potential regulatory roles of non-coding variants with nearby genes.
